# Selective prediction for extracting unstructured clinical data

**DOI:** 10.1101/2022.11.15.22282368

**Authors:** Akshay Swaminathan, Ivan Lopez, William Wang, Ujwal Srivastava, Edward Tran, Aarohi Bhargava-Shah, Janet Y Wu, Alexander Ren, Kaitlin Caoili, Brandon Bui, Layth Alkhani, Susan Lee, Nathan Mohit, Noel Seo, Nicholas Macedo, Winson Cheng, Charles Liu, Reena Thomas, Jonathan H. Chen, Olivier Gevaert

## Abstract

Electronic health records represent a large data source for outcomes research, but the majority of EHR data is unstructured (e.g. free text of clinical notes) and not conducive to computational methods. While there are currently approaches to handle unstructured data, such as manual abstraction, structured proxy variables, and model-assisted abstraction, these methods are time-consuming, not scalable, and require clinical domain expertise. This paper aims to determine whether selective prediction, which gives a model the option to abstain from generating a prediction, can improve the accuracy and efficiency of unstructured clinical data abstraction. We trained selective prediction models to identify the presence of four distinct clinical variables in free-text pathology reports: primary cancer diagnosis of glioblastoma (GBM, n = 659), resection of rectal adenocarcinoma (RRA, n = 601), and two procedures for resection of rectal adenocarcinoma: abdominoperineal resection (APR, n = 601) and low anterior resection (LAR, n = 601). Data were manually abstracted from pathology reports and used to train L1-regularized logistic regression models using term-frequency-inverse-document-frequency features. Data points that the model was unable to predict with high certainty were manually abstracted. All four selective prediction models achieved a test-set sensitivity, specificity, positive predictive value, and negative predictive value above 0.91. The use of selective prediction led to sizable gains in automation (anywhere from 57% to 95% reduction in manual abstraction of charts across the four outcomes). For our GBM classifier, the selective prediction model saw improvements to sensitivity (0.94 to 0.96), specificity (0.79 to 0.96), PPV (0.89 to 0.98), and NPV (0.88 to 0.91) when compared to a non-selective classifier. Selective prediction using utility-based probability thresholds can facilitate unstructured data extraction by giving “easy” charts to a model and “hard” charts to human abstractors, thus increasing efficiency while maintaining or improving accuracy.

## INTRODUCTION

Electronic health records (EHRs) are a valuable data source for clinical outcomes research and quality improvement^1,2^. EHRs typically contain structured and unstructured data. Structured data refers to data captured from drop-down menus, multi-select menus, or other data modalities that follow a consistent format when entered into the EHR^3^. Unstructured data refers to free-form text, prose, or data that does not follow a consistent format when entered into the EHR. While structured data like medication orders and ICD (International Classification of Diseases) codes have been used for research purposes, important variables for outcomes research (e.g. diagnosis dates, cancer progression and recurrence events, treatment response, comorbidities, adverse events, mortality, etc.) are typically available only as unstructured data. It has been estimated that over 80% of all healthcare data is unstructured^4^ — therefore, we need scalable methods of extracting unstructured data to enable clinical research and quality improvement efforts.

Unstructured variables in EHRs are usually dealt with using manual abstraction and proxy variables available as structured data^5^. Manual abstraction is time-consuming and not scalable, especially with the increase in EHR data^6^. Additionally, manual abstraction can be expensive if a highly trained professional (e.g. clinician) is required to comprehend the clinical nuances of EHR data.

In cases where manual abstraction is logistically infeasible, structured data proxy variables have been used in population-based research. Crucially, structured proxies are imperfect and can introduce bias into downstream analyses. For example, the presence of an ICD code (structured) may be used as a proxy for clinician-confirmed diagnosis (unstructured) for a certain condition. However, the use and accuracy of ICD codes may vary substantially across sites, is not validated, and can present challenges when updating to newer versions^7–9^. Additionally, CPT codes are frequently miscoded^10^.

Given these limitations of manual clinical abstraction, new approaches that leverage machine learning to increase efficiency and accuracy must be explored^6,11^. Despite recent advances in the field, machine learning (ML) models still do not perform as well as humans on several unstructured data extraction tasks, such as those involving negation terms or small misspellings, and tasks requiring reasoning from context^12–15^. Therefore, for use cases that require high accuracy, model-based abstraction may not always be acceptable.

Another approach to unstructured data extraction is model-assisted abstraction, where humans use the output of machine learning models to improve the efficiency or accuracy of manual abstraction^16^. One common approach for model-assisted abstraction is using models to identify records that may require human review^16^. For example, this may be used to build a cohort of patients that meet certain inclusion criteria. While this approach may improve abstraction efficiency, charts not flagged by the model may never be included in the final dataset, leading to lower sample sizes and systematic exclusion of certain patients. Therefore, there is still no effective way to synthesize the terabytes of unstructured EHR data produced daily, and as a result, the quality and quantity of clinical outcomes research are limited^17^.

To address the above limitations of manual abstraction and model-assisted abstraction, we used selective prediction, a modeling framework that allows a model to abstain from generating a prediction under certain scenarios, for example when uncertainty in the prediction is high. We hypothesized that this approach would lead to improved accuracy compared to traditional prediction for data points where the model generated a prediction. We applied this workflow to four clinical variables: diagnosis of glioblastoma (GBM) (binary, yes/no), diagnosis of rectal adenocarcinoma (binary, yes/no), laparoscopic surgical resection of colorectal adenocarcinoma (binary, yes/no), laparoscopic surgical resection approach for colorectal adenocarcinoma (categorical, LAR/APR/other). The final output of our models is high-quality structured data that can be used for research or quality improvement.

## METHODS

### Approach

First, manual abstraction of a fixed number of data points was performed by human abstractors (Figure 1a). This decreases the costs typically associated with having highly trained clinicians perform chart abstraction. Second, natural language processing (NLP) selective prediction models were developed using the data collected by manual abstraction. In contrast to typical ML models that generate a prediction for any input, selective prediction models may abstain from generating a prediction for certain data points (Figure 1b)^18^. Third, we leveraged selective prediction modeling to generate predictions for data points that the model can predict with high accuracy and used manual abstraction for data points that the model abstains from predicting. Lastly, we applied a series of quality checks to ensure the fidelity of the final variables.

**Fig. 1a.**
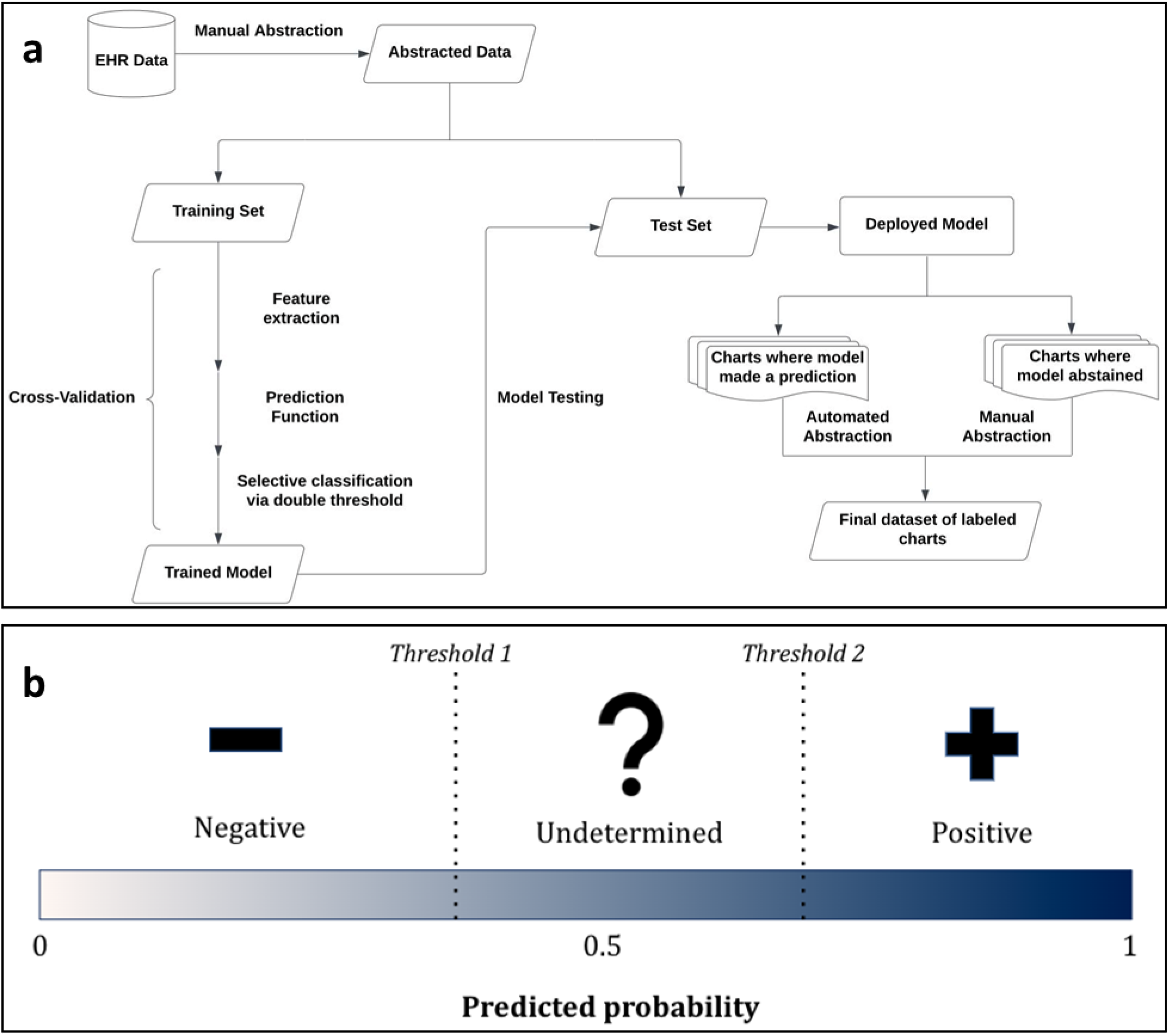
Diagram depicting the modeling workflow used to extract unstructured variables. EHR data is abstracted, then used to train and test a model that either predicts an outcome or abstains from a prediction. Charts for which a prediction is not generated are abstracted manually. **b** Depiction of selective prediction via utility-based thresholding. Two thresholds (Threshold 1 and Threshold 2) are applied to a continuous predicted probability to yield three possible classifications: negative (predicted probability < Threshold 1), positive (predicted probability > Threshold 2), and undetermined (Threshold 1 ≤ predicted probability ≤ Threshold 2). Thresholds are selected based on real world utilities so as to minimize the total cost, defined as (# of FP) × (cost of FP) + (# of FN) × (cost of FN) + (# of undetermined) × (cost of undetermined), where # = number, FP = false positive, and FN = false negative.

## Data source

This study was a retrospective analysis of EHR data collected at Stanford Hospital, Stanford, CA. This data source included any patient treated at Stanford from 1998-2022. Ethics approval was granted through Stanford University IRB (#50031).

## Study Population

### Glioblastoma cohort

Patients with mention of the word “glioblastoma” in a pathology report were included. Filtering for pathology reports with the word “glioblastoma” is an effective way of identifying patients who are likely to have primary GBM, since any patient diagnosed with primary GBM would have a pathology report indicating that diagnosis (high sensitivity). Patients younger than 18 at the date of the pathology report that contained the word “glioblastoma” were excluded due to differences in the clinical management of pediatric GBM. The remaining 1195 patients were considered for subsequent analyses. Of these, 629 patients (659 charts) were randomly selected for manual abstraction.

### Colorectal cancer cohort

The colorectal cancer cohort was created by selecting patients between January 2012 and December 2021 that had ICD-9 and ICD-10 codes for rectal cancer (154.0 and 154.1 for IDC-9 and C19 and C20 for ICD-10), and Current Procedural Terminology (CPT) codes for treatment of rectal cancer (45110, 45111, 45112, 45114, 45116, 45126, 45395, 44146, 44208, 48.4, 48.5, 48.6, 1007599, 1007604). Patients younger than 18 at the date of the pathology report were excluded due to differences in clinical management. Of the remaining 726 patients, 298 patients (601 charts) were randomly selected for manual abstraction and considered for subsequent analysis.

## Outcomes

### Confirmation of primary glioblastoma diagnosis

Primary glioblastoma diagnosis was determined by reviewing the relevant pathology reports of patients with suspected GBM. In general, patients diagnosed via surgical resection, biopsy, or specimen review with GBM (WHO grade IV), with or without oligodendroglial or gliosarcoma features, were considered to have primary glioblastoma. Patients with transformed or recurrent GBM diagnoses were not considered to have primary glioblastoma.

### Confirmation of rectal adenocarcinoma diagnosis

Rectal adenocarcinoma diagnosis was determined by reviewing the relevant pathology reports of patients with suspected rectal adenocarcinoma. In general, patients diagnosed via surgical resection of rectal adenocarcinoma were considered to have a rectal adenocarcinoma diagnosis.

### Confirmation of laparoscopic surgical resection of rectal adenocarcinoma

Confirmation of laparoscopic surgical resection (binary) of rectal adenocarcinoma was made after a patient was confirmed to have been diagnosed with rectal adenocarcinoma. There are two laparoscopic surgical approaches for resection of rectal adenocarcinoma of interest. The first approach is low anterior resection (LAR), which involves removing part of the rectum. The second approach is abdominoperineal resection (APR), which involves removing the anus, rectum, and sigmoid colon. A third, less common approach, is called pelvic exenteration (PE), which involves removing reproductive organs, the bladder or rectum or both, and lymph nodes in the pelvis. Here, we built models to identify patients who underwent APR or LAR. A model to identify patients who underwent PE was not built due to insufficient data.

## Data abstraction

Of the patients that met the initial inclusion criteria, 629 patients were randomly selected for manual abstraction to collect training data for the NLP models. To confirm GBM diagnosis, we used the methodology described below to perform abstraction. For the variables related to colorectal cancer, we used existing data that had been manually abstracted for other research purposes.

First, a chart review was performed to understand how GBM diagnosis was documented in the EHR. Second, based on the findings from the preliminary chart review, a detailed abstraction instruction guide was created that described how to determine a diagnosis of GBM from a patient’s chart. Instructions for dealing with ambiguous cases (ex. gliosarcoma or glioblastoma with oligodendroglioma features) were included, as well as guidance for cases that the abstractor could not determine. Third, a standardized abstraction data entry tool was developed, and a pilot round of abstraction was conducted to refine the instruction guide and data entry tool. The data entry tool had a “flag” feature that abstractors could use to indicate charts for which they were not able to make a determination. Fourth, patient charts were allocated to non-clinician abstractors (with duplication to allow for calculation of inter-rater reliability) and abstraction was performed. A subset of charts was assigned to clinician abstractors to calculate the accuracy of non-clinician abstractors. Lastly, all flagged charts were reviewed, and data quality checks were performed.

We ran selective prediction models on 4 datasets taken from pathology reports, each of which confirmed GBM diagnosis, resection of rectal adenocarcinoma (RRA), LAR of rectal adenocarcinoma, or APR of rectal adenocarcinoma (Table 1).

**Table 1.** Questions and summary of responses for the cost survey (n = 6).

| Common scenario:<br>Suppose you have a group of patients, each with one medical chart. You need to decide whether each of these patients should be included in a dataset containing patients with GBM. This dataset will be used for outcomes research and quality improvement efforts.<br>Since no prediction model is 100% accurate, it is expected that it will make some mistakes in its decision to include/exclude a patient from the dataset. Alternatively, you can manually review a patient's case and make the right decision with absolute certainty, but note that manually reviewing the chart is time consuming. |  |  |  |
| --- | --- | --- | --- |
| Question | Average | Minimum | Maximum |
| How many patient charts would you be willing to review manually to prevent one patient who is healthy from being incorrectly included into the GBM dataset (i.e. false positive)?* | 13.5 | 10 | 20 |
| How many patient charts would you be willing to review manually to prevent one patient who has GBM from being left out of the dataset (i.e. false negative)?* | 8 | 3 | 15 |

## Statistical Analysis

### Feature engineering and feature selection

For each variable, modeling was performed at the level of a patient chart, and each patient may have multiple charts. A randomly selected 60% of charts were used for model training, and the remaining 40% were used for model testing. Features were derived entirely from the free text of pathology reports. Using the text of these documents, TF-IDF (term-frequency-inverse-document-frequency) scores were calculated for unigrams, bigrams, trigrams, and 4-grams. Briefly, TF-IDF is a method used to quantify the relative frequency of an n-gram (an n-word phrase) in a corpus of documents. For example, the TF-IDF score for the unigram “cancer” in a patient’s pathology report is calculated by dividing the frequency of the word “cancer” in that patient’s pathology report (term frequency) by the frequency of the word “cancer” across all pathology reports across all patients (inverse document frequency). L1-regularized logistic regression (Lasso) was used for feature selection and prediction. Hyperparameters (regularization for Lasso and sparsity for document term matrix) were tuned using 10-fold cross-validation. The metric for cross-validation was misclassification cost (described below). Selected features were examined for sensibility and clinical meaningfulness (Supplementary Table 1).

### Binary classification

For each binary variable, the output of the lasso regression model was a probability between 0 and 1 indicating the probability that the given chart contains the variable of interest. Typically, a probability threshold is applied to convert a probability prediction model into a classifier. In selecting an optimal probability threshold, relative costs for false negatives and false positives must be specified. Here, “cost” refers not to monetary cost but rather real-world utilities. In the example of GBM diagnosis, the cost of a false positive corresponds to the cost of incorrectly labeling a chart as having GBM, and the cost of a false negative corresponds to the cost of incorrectly labeling a chart as not having GBM. The former results in incorrectly including a patient in the final dataset, whereas the latter results in incorrectly excluding a patient from the final dataset.

We implemented selective prediction by selecting two probability thresholds instead of one. Any chart with a predicted probability of falling between the two thresholds is considered “undetermined”. The idea behind this approach is that a predicted probability very close to 0 or 1 indicates higher model confidence. By labeling charts whose predicted probability is far from 0 or 1 as “undetermined”, the model performance on the charts not labeled “undetermined” is likely to be better than the model performance on the overall sample.

The double threshold requires specifying a relative cost to labeling a chart as “undermined”, as well as costs for false positives and false negatives. A grid search was used to identify the two thresholds which minimize the total cost, defined as follows:

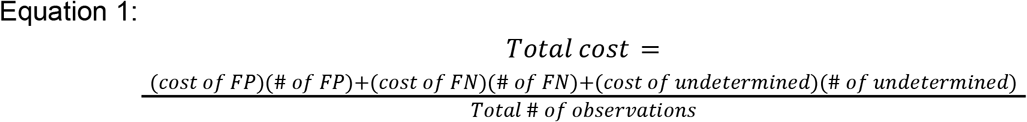

where, # is number, FP is false positive, and FN is false negative. With this double threshold approach, we can expect abstraction efficiency gains equal to 100% minus the “undetermined rate”, or the percentage of charts labeled as “undetermined”. Although the model may not be able to classify 100% of charts, we expect more accurate performance on the subset of charts the model is able to classify, thus leading to gains in abstraction efficiency with minimal loss of accuracy.

We compare the double threshold approach to the typical single threshold approach, where predicted probabilities above or below the threshold are labeled “positive” or “negative” respectively.

To quantify the relative costs of FPs, FNs, and undetermined as viewed by researchers (the end users of the model), we created a survey that uses a regret-based approach^19^. This validated method asks respondents to consider their preferences and willingness to accept tradeoffs in hypothetical scenarios about patient care. Our survey presents a scenario with 100 patient charts to be classified by an imperfect prediction model and asks respondents to quantify how many patient charts they would be willing to review manually to avoid a false positive and false negative. The responses can be used to generate cost ratios for FP:undetermined and FN:undetermined, respectively. A total of seven clinician and abstractor responses were received. Responses were scanned for consistency and follow up conversations were used to understand motivations for all responses. After removing one outlier response, the remaining responses had similar values and reasoning. A simple average of the remaining six preferences gave the cost ratio for FP:FN:undetermined of 13.5:8:1. This means that the model returning a false positive is 13.5 times more costly than abstaining. Similarly, a false negative result is 8 times more costly than the model abstaining.

### Model evaluation

Misclassification cost, sensitivity, specificity, positive predictive value (PPV), and negative predictive value (NPV) were calculated, treating the abstracted labels as ground truth. In addition, we introduce three metrics for the evaluation of selective prediction models: undetermined rate, positive undetermined rate, and negative undetermined rate. Positive undetermined rate is defined as the number of positive observations labeled as undetermined (undetermined positives or UP) over the total number of positive observations within the training dataset (TP + FN + UP). Similarly, the negative undetermined rate is defined as the number of negative observations (UN) labeled as undetermined over the total number of negative observations within the training dataset (TN + FP + UN). Positive and negative undetermined rates give insight into the composition of the undetermined rate. The undetermined rate is defined as the number of predictions a selective prediction model abstains from making (UN + UP), over the total number of possible predictions (UN + UP + FN + FP + TN + TP). A selective prediction model with a lower undetermined rate yields greater efficiency gains than a model with a larger undetermined rate and similar levels of accuracy. For example, if there is an equal proportion of cases and controls in the training dataset, but the positive undetermined rate is much larger than the negative undetermined rate, this suggests the selective prediction model is struggling to predict labels for cases accurately.

After training, the final model was applied to all unlabeled data, yielding three types of predictions: “positive”, “negative” or “undetermined”. For charts labeled positive or negative, the model’s prediction was considered final. Charts labeled “undetermined” were those for which the model abstained from making a definitive prediction. These charts were given to abstractors for manual review.

### Failure analysis

Failure analysis involved a manual review of observations where the model prediction resulted in false positives or false negatives during cross-validation. The goal of failure analysis was to determine the reasons for the model’s incorrect prediction. In some cases, the model’s prediction was correct and erroneously labeled as a false positive or false negative due to abstractor error. Potential reasons for abstractor error include mixing up dates of records, misreading or misinterpreting similar words or synonyms, and failing to notice words and phrases. In these cases, the underlying data was amended, the model was retrained, and model accuracy metrics were recalculated.

### Comparison to structured data proxies

We assessed whether the output of selective prediction models for GBM diagnosis was more accurate than EHR-derived structured proxy variables. This analysis was not performed for the colorectal cancer cohorts because structured variables were considered in the inclusion criteria for those cohorts. Structured proxy variables for diagnosis of GBM may include the presence of an ICD or CPT code or documented treatment with antineoplastics typically used to treat GBM. The following ICD codes relevant to GBM were included: C71.0 -C71.9 and C72.9 in the ICD-10 scheme; C191.9 in the ICD-9 scheme^20–22^. CPT codes related to “glioblastoma” (81287, 81345), “malignant neoplasm of the brain” (70554, 70555, 78600, 78601, 78605, 78606, 78608, 78609, 78610), and surgical procedures related to GBM (61510) were included^23^. The proxy for treatment included any documented order or administration of temozolomide or bevacizumab. The labels generated by the model were compared with these structured data elements using the performance metrics described above to quantify the increased accuracy obtained by using an NLP-based model.

## RESULTS

### Cost survey

Based on the survey conducted (n = 6), a cost ratio for FP:FN:undetermined was assigned as 13.5:8:1 (Table 1).

### Colorectal cancer models

Overall, our double threshold models performed better than the single threshold models, with 16%, 35%, and 41% decreases in total cost compared to single threshold models for resection of rectal adenocarcinoma (RRA), abdominoperineal resection (APR), and low anterior resection (LAR), respectively (Table 2). Among the three colorectal cancer (CRC) models, the LAR double threshold classifier had the highest undetermined rate (10%), where 90% of charts were automatically labeled by the LAR classifier. This led to improvements over single threshold modeling in sensitivity (0.71 to 0.91), specificity (0.99 to 1.00), PPV (0.94 to 0.98), and NPV (0.94 to 0.99). Our RRA double threshold classifier had the second highest undetermined rate (9%), where 91% of charts were automatically labeled by the RRA classifier. Similarly, we saw improvements over single threshold modeling in sensitivity (0.87 to 0.93), specificity (0.92 to 0.93), PPV (0.90 to 0.92), and NPV (0.90 to 0.94). Lastly, our APR double threshold classifier had the lowest undetermined rate (5%), where 95% of charts were automatically labeled by our APR classifier. This led to improvements over single threshold modeling in sensitivity (0.81 to 0.91), specificity (0.99 to 0.99), PPV (0.88 to 0.94), and NPV (0.98 to 0.99).

**Table 2:**
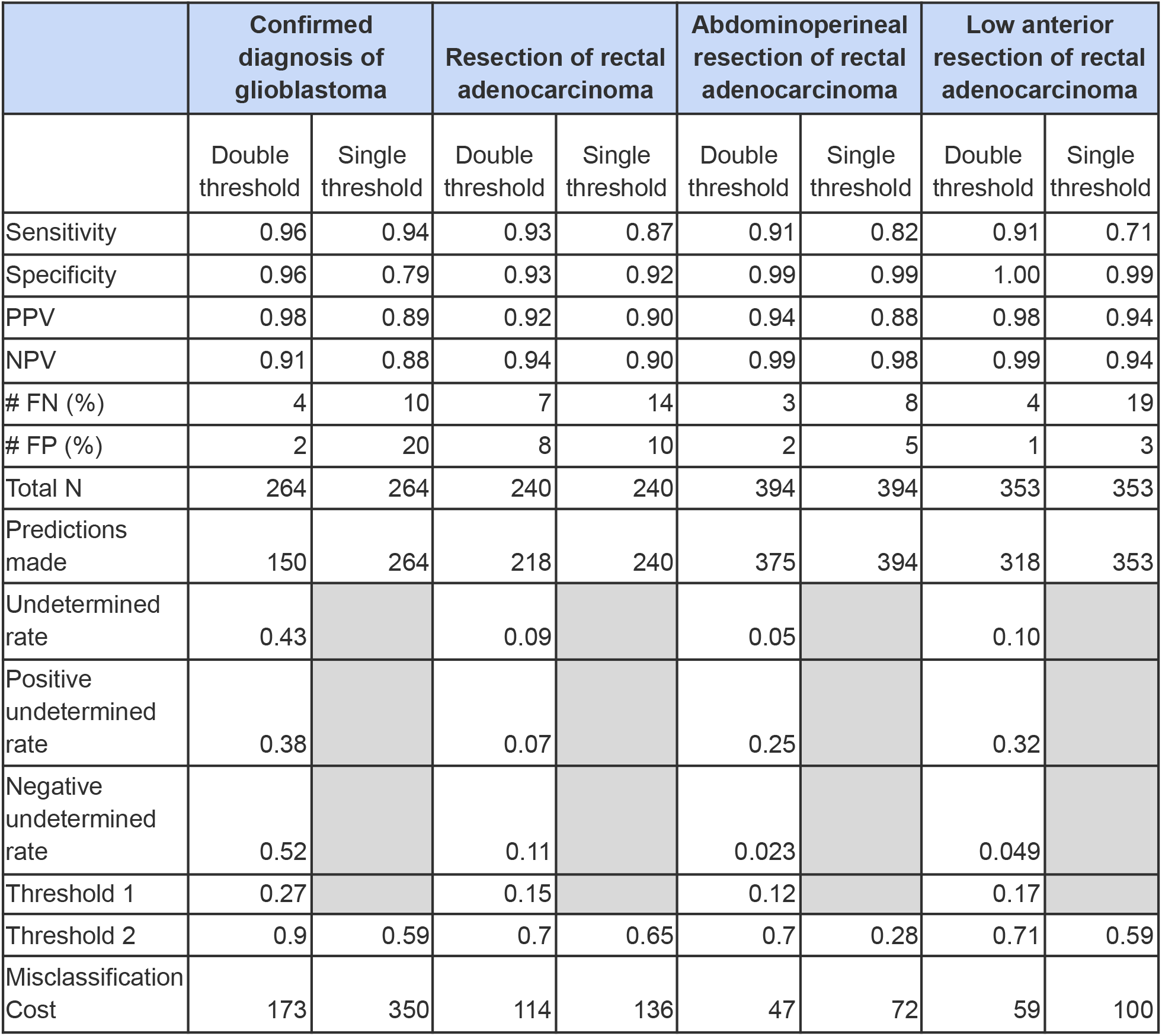
Test set performance of classification models (single threshold and double threshold) for confirmed diagnosis of glioblastoma, resection of rectal adenocarcinoma, abdominoperineal resection of rectal adenocarcinoma, and low anterior resection of rectal adenocarcinoma. (note: positive undetermined rate = UP/(TP+FN+UP) and negative undetermined rate = UN/(TN+FP+UN), where UP (undetermined positives) is the number of positive observations labeled as undetermined, TP+FN+UP represents the total number of positive observations in the training dataset, UN (undetermined negatives) is the number of negative observations labeled as undetermined, and TN+FP+UN represents the total number of negative observations in the training dataset).

### Glioblastoma model

The double threshold model performed better than the single threshold models, with a 51% decrease in total cost compared to the single threshold models (Table 2). The GBM double threshold classifier had an undetermined rate of 43%, meaning that 57% of charts were automatically labeled by the model. This led to improvements over single threshold modeling in sensitivity (0.94 to 0.96), specificity (0.79 to 0.96), PPV (0.89 to 0.98), and NPV (0.88 to 0.91).

For GBM, the double threshold model outperformed all structured proxy variables, yielding costs anywhere from 84.8% to 87.1% lower than those of structured proxy variables (Table 3).

**Table 3.** Performance metrics of structured proxy variables (ICD codes, CPT codes, and GBM-related medication) in predicting confirmed diagnosis of GBM.

| Metric | Any GBM-related ICD Code | Any GBM-related CPT Code | Received GBM-related medication at any time |
| --- | --- | --- | --- |
| Sensitivity | 0.62 | 0.36 | 0.21 |
| Specificity | 0.46 | 0.59 | 0.84 |
| PPV | 0.70 | 0.64 | 0.71 |
| NPV | 0.38 | 0.32 | 0.34 |
| # FP | 46 | 35 | 14 |
| # FN | 65 | 108 | 135 |
| Misclassification cost | 1141 | 1337 | 1269 |

Compared to ICD codes, the double threshold model had higher sensitivity (0.96 to 0.62), specificity (0.96 to 0.46), PPV (0.98 to 0.70) and NPV (0.91 to 0.38). Compared to CPT codes, the double threshold model had higher sensitivity (0.96 to 0.36), specificity (0.96 to 0.59), PPV (0.98 to 0.64) and NPV (0.91 to 0.32). Compared to the proxy variable of GBM-related treatment, the double threshold model had higher sensitivity (0.96 to 0.21), specificity (0.96 to 0.84), PPV (0.98 to 0.71) and NPV (0.91 to 0.34).

## DISCUSSION

EHR data is a valuable resource for investigating clinical questions, but it is hard to analyze in its unstructured form. Currently, data extraction techniques rely on manual abstractors, which can be time-consuming and resource-intensive. We propose an approach that leverages selective prediction and human abstractors to improve the accuracy and efficiency of clinical data abstraction. Specifically, we used selective prediction implemented via cost-based probability thresholding to allow our classifier to abstain from predicting certain data points, which were then given to a human for manual abstraction.

The double threshold model led to sizable gains in automation (anywhere from 57% to 95% across our four outcomes) and for GBM diagnosis, outperformed all structured proxy variables in terms of misclassification cost.

For any prediction model, the resulting prediction has a higher chance of being wrong when model uncertainty is high. In many healthcare settings, making a wrong prediction is more harmful than not making a prediction at all. Selective prediction has the potential to improve efficiency and accuracy in healthcare applications where the cost of abstaining from a prediction is lower than the cost of a misclassification. One such scenario is automated chart abstraction for cohort selection because the cost of abstaining is the cost of manually reviewing a chart. The relative preference for abstaining compared to making an incorrect classification is reflected in the results of the cost survey. Other examples of selective prediction exist; for example, Kotropoulos and Arce reported building a linear classifier with the option to postpone decision-making, which they called a rejection^24,25^. In this paper, postponed decisions required further advice by a domain expert in the context of the paper’s intended application. Another example comes from Guan et al., where they used a selective prediction classification model to abstain from prediction when the classification task has a large amount of uncertainty^26^. The results for our double-threshold model are comparable to state-of-the-art algorithms; for instance, one set of models for various clinical concepts reported an average sensitivity of 95.5% and PPV of 95.3% for unstructured data^27^.

Defining misclassification cost is a critical step in developing classification models. Common metrics for assessing ML model performance include F1 score, area under the curve (AUC), and calibration, but such metrics fail to consider real-world consequences of model errors^28^. These numbers have little context in the clinical setting e.g. there is no cutoff F1 score that indicates a model is safe to use in making clinical decisions. Assigning costs for false positives and false negatives is an attempt to capture the difficulty of trade-offs in medical decision-making and tie the output of the model to the would-be impact of the model’s output. To establish cost ratios, we used stakeholder surveys but other methods may be implemented.

Through this survey, stakeholders must implicitly consider how much they value quantities such as clinical accuracy, time, and monetary cost. Since there is no single right cost ratio, a survey that directly asks relevant stakeholders about their preferences, as we did, can help elicit the relative costs associated with the model outputs. Such a survey that makes people consider consequences and anticipate regret has been suggested to be effective because it combines both intuitive and deliberative systems of thinking^19^.

One key question that selective prediction raises is how to deal with the data points where the model abstained. One simple approach is to have humans review all undetermined data points by hand. This may be preferable when the number of undetermined data points is small, when the total cost of manual review is low, or when the outcome is difficult to predict for undetermined data points. Beyond a certain number of training data points, the signal-to-noise ratio decreases and improvements in classification performance diminish^29^. The manual review of undetermined data points could lend insight into methods to help fine-tune models. Another approach could leverage active learning techniques like uncertainty sampling to preferentially sample undetermined data points and retraining on charts that were initially labeled as uncertain.

Our approach has some limitations. First, we tested the algorithm on clinical oncology documents (pathology reports) and binary variables (diagnosis and procedures). Further testing is needed to confirm that the model can work on a variety of different datasets and variables.

Second, selective prediction could theoretically result in a significant portion of undetermined data points without substantial increases in accuracy. Adjusting the cost ratios may ameliorate this. As the cost assigned to an undetermined increases, the model will make more predictions, but one faces the trade off of more false positives and false negatives in this case. Third, we use predicted probabilities as proxies for model uncertainty, which may not be the optimal metric for thresholding. In the active learning literature, alternative metrics for quantifying model uncertainty have been proposed, including Shannon entropy and variance in query-by-committee predictions^30^.

Selective prediction using cost-based probability thresholding can semi-automate unstructured EHR data extraction by giving “easy” charts to a model and “hard” charts to human abstractors, thus increasing efficiency while maintaining or improving accuracy. A semi-automated abstraction workflow substantially outperforms structured proxy variables like ICD codes on a binary classification task, generating higher quality datasets that can be used for outcomes research.

## Data Availability

All data needed to evaluate the conclusions are present in the paper and in the Supplementary Materials. The datasets generated analyzed during the current study are not publicly available due to patient privacy but are available from the corresponding author (OG) on reasonable request.

## AUTHOR CONTRIBUTIONS

AS, IL, and OG conceived the idea. AS and IL performed data verification. AS, IL, WW, ET, US, ABS, JW, AR, BB, LA, SL, NM, NS, NM, WC, and CL carried out data acquisition. IL, AS, WW, US, and ET performed data analysis and interpretation. IL, AS, WW, US, and ET wrote the draft of the manuscript and made critical revisions. OG, JHC, and RT supervised the project. All authors have read and approved the manuscript.

## Supplementary information

**Supplementary Table 1:** This table contains the top 10 features selected from each of our 4 classifiers. Features are listed in descending order of importance with the feature at the top selected as the most important feature for that classifier.

| Top 10 Features Selected |  |  |  |
| --- | --- | --- | --- |
| Confirmed diagnosis of glioblastoma | Resection of rectal adenocarcinoma | Abdominoperineal resection of rectal adenocarcinoma | Low anterior resection of rectal adenocarcinoma |
| – glioblastoma | – invasive adenocarcinoma | anterior resection – | adipose tissue. |
| – glioblastoma (who | ( / / | anterior resection tumor | and distal |
| – glioblastoma, | (cm) negative | operation: laparoscopic | anterior resection |
| hospital | adenocarcinoma (see | sigmoid colon | anterior resection – |
| is frozen | distal | specimen labeled<br>“proximal | anterior resection tumor |
| mitotic | line | tumor regression | loop |
| necrosis are | mucosa | circumferential | operation: laparoscopic |
| tumor cells | opened | fragment of | rectal cancer operation: |
| discussed | possible | low anterior | sigmoid |
| grade iv | rectal cancer operation: | muscularis propria | (cm) negative |

**Supplementary Table 2.** Areas under the receiver operator curve (AUC) and calibration slope and intercept for glioblastoma (GBM), resection of rectal adenocarcinoma (RRA), abdominoperineal resection (APR), and low anterior resection (LAR) traditional (non-selective prediction) models.

| Model | AUC | Calibration slope | Calibration intercept |
| --- | --- | --- | --- |
| GBM | 0.93 | 1.22 | -0.38 |
| RRA | 0.94 | 0.52 | -0.32 |
| APR | 0.99 | 1.36 | -0.01 |
| LAR | 0.98 | 1.48 | 0.31 |

**Supplementary Table 3.** Results of model failure analysis for the four NLP double threshold models. False positives (FP) and false negatives (FN) were manually reviewed to determine whether they were due to model error or abstractor error.

| Metric | Confirmed diagnosis of glioblastoma | Confirmed diagnosis of rectal adenocarcinoma | Low anterior resection of rectal adenocarcinoma | Abdominoperineal resection of rectal adenocarcinoma |
| --- | --- | --- | --- | --- |
| FPs due to model error | 0 | 4 (50% of all FPs) | 1 (100% of all FPs) | 2 (100% of all FPs) |
| FPs due to abstractor error | 0 | 4 (50% of all FPs) | 0 | 0 |
| FNs due to model error | 0 | 7 (100% of all FNs) | 2 (50% of all FNs) | 2 (67% of all FNs) |
| FNs due to abstractor error | 0 | 0 | 2 (50% of all FNs) | 1 (33% of all FNs) |

**Supplementary Figure 1.**
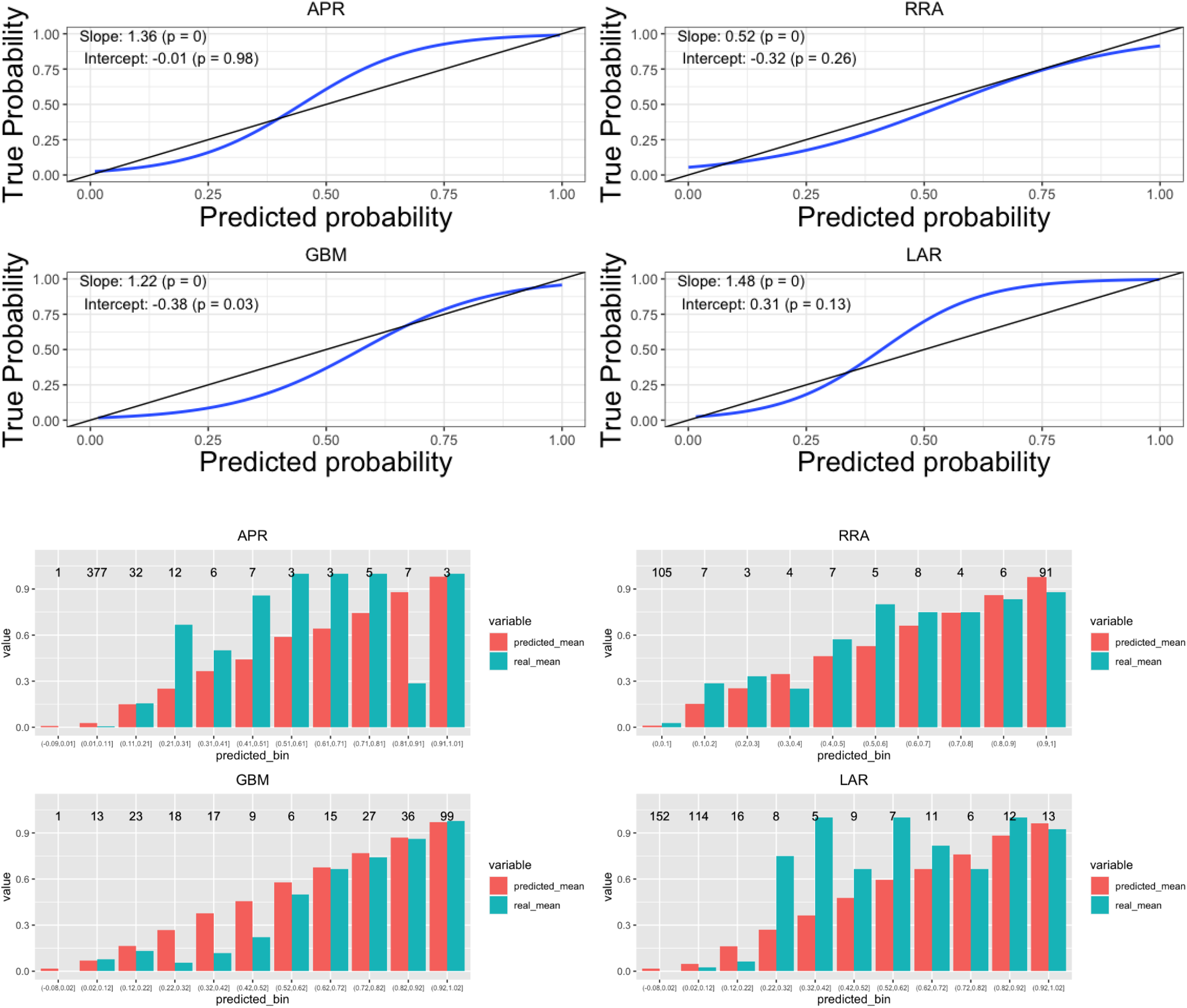
Calibration metrics for glioblastoma (GBM), resection of rectal adenocarcinoma (RRA), abdominoperineal resection (APR), and low anterior resection (LAR) traditional (non-selective prediction) models

## References

1 Improved Diagnostics & Patient Outcomes | HealthIT.gov. https://www.healthit.gov/topic/health-it-and-health-information-exchange-basics/improved-diagnostics-p atient-outcomes (accessed Sept 14, 2022).

2 Hecht J. The future of electronic health records. Nature 2019; 573: S114–6.

3 Polnaszek B, Gilmore-Bykovskyi A, Hovanes M, et al. Overcoming the Challenges of Unstructured Data in Multi-site, Electronic Medical Record-based Abstraction. Med Care 2016; 54: e65–72.

4 Kong H-J. Managing Unstructured Big Data in Healthcare System. Healthc Inform Res 2019; 25: 1–2.

5 Yang S, Varghese P, Stephenson E, Tu K, Gronsbell J. Machine learning approaches for electronic health records phenotyping: A methodical review. 2022; DOI:10.1101/2022.04.23.22274218.

6 Alzu‘bi AA, Watzlaf VJM, Sheridan P. Electronic Health Record (EHR) Abstraction. Perspect Health Inf Manag 2021; 18: 1g.

7 Kaur R, Ginige JA, Obst O. A Systematic Literature Review of Automated ICD Coding and Classification Systems using Discharge Summaries. 2021; published online July 12. DOI:10.48550/arXiv.2107.10652

8 Rasmy L, Tiryaki F, Zhou Y, et al. Representation of EHR data for predictive modeling: a comparison between UMLS and other terminologies. J Am Med Inform Assoc 2020; 27: 1593–9.

9 O‘Malley KJ, Cook KF, Price MD, Wildes KR, Hurdle JF, Ashton CM. Measuring Diagnoses: ICD Code Accuracy. Health Serv Res 2005; 40: 1620–39.

10 King MS, Sharp L, Lipsky MS. Accuracy of CPT Evaluation and Management Coding by Family Physicians. The Journal of the American Board of Family Practice 2000; 14: 184–92.

11 Bommasani R, Hudson DA, Adeli E, et al. On the Opportunities and Risks of Foundation Models. 2022; published online July 12. DOI:10.48550/arXiv.2108.07258.

12 Wu S, Roberts K, Datta S, et al. Deep learning in clinical natural language processing: a methodical review. Journal of the American Medical Informatics Association 2020; 27: 457–70.

13 Lin BY, Gao W, Yan J, Moreno R, Ren X. RockNER: A Simple Method to Create Adversarial Examples for Evaluating the Robustness of Named Entity Recognition Models. 2021; published online September 12. DOI:10.48550/arXiv.2109.05620.

14 Pruthi D, Dhingra B, Lipton ZC. Combating Adversarial Misspellings with Robust Word Recognition. 2019; published online August 29. DOI:10.48550/arXiv.1905.11268.

15 Singh PK, Paul S. Deep Learning Approach for Negation Handling in Sentiment Analysis. IEEE Access 2021; 9: 102579–92.

16 Birnbaum B, Nussbaum N, Seidl-Rathkopf K, et al. Model-assisted cohort selection with bias analysis for generating large-scale cohorts from the EHR for oncology research. 2020; published online January 13. http://arxiv.org/abs/2001.09765 (accessed September 14, 2022).

17 Botsis T, Hartvigsen G, Chen F, Weng C. Secondary Use of EHR: Data Quality Issues and Informatics Opportunities. Summit on Translat Bioinforma 2010; 2010: 1–5.

18 Xin J, Tang R, Yu Y, Lin J. The Art of Abstention: Selective Prediction and Error Regularization for Natural Language Processing. In: Proceedings of the 59th Annual Meeting of the Association for Computational Linguistics and the 11th International Joint Conference on Natural Language Processing (Volume 1: Long Papers). Online: Association for Computational Linguistics, 2021: 1040–51.

19 Tsalatsanis A, Hozo I, Vickers A, Djulbegovic B. A regret theory approach to decision curve analysis: A novel method for eliciting decision makers‘ preferences and decision-making. BMC Med Inform Decis Mak 2010; 10: 51.

20 2021/2022 ICD-10-CM Index > ‘Glioblastoma‘. https://www.icd10data.com/ICD10CM/Index/G/Glioblastoma (accessed September 14, 2022).

21 2022 ICD-10-CM Codes C72*: Malignant neoplasm of spinal cord, cranial nerves and other parts of central nervous system. https://www.icd10data.com/ICD10CM/Codes/C00-D49/C69-C72/C72 -(accessed September 14, 2022).

22 2022 ICD-10-CM Codes C71*: Malignant neoplasm of brain. https://www.icd10data.com/ICD10CM/Codes/C00-D49/C69-C72/C71 - (accessed September 14, 2022).

23 Medical Billing Codes Search - CPT, ICD 9, ICD 10 HCPCS Codes & Articles, Guidelines Codify by AAPC. https://www.aapc.com/codes/code-search/ (accessed September 14, 2022).

24 Kompa B, Snoek J, Beam AL. Second opinion needed: communicating uncertainty in medical machine learning. npj Digit Med 2021; 4: 1–6.

25 Kotropoulos C, Arce GR. Linear Classifier with Reject Option for the Detection of Vocal Fold Paralysis and Vocal Fold Edema. EURASIP J Adv Signal Process 2009; 2009: 1–13.

26 Guan H, Zhang Y, Cheng HD, Tang X. Bounded-abstaining classification for breast tumors in imbalanced ultrasound images. International Journal of Applied Mathematics and Computer Science 2020; 30: 325–36.

27 Hernandez-Boussard T, Monda KL, Crespo BC, Riskin D. Real world evidence in cardiovascular medicine: ensuring data validity in electronic health record-based studies. Journal of the American Medical Informatics Association 2019; 26: 1189–94.

28 Vickers AJ, Calster BV, Steyerberg EW. Net benefit approaches to the evaluation of prediction models, molecular markers, and diagnostic tests. BMJ 2016; 352: i6.

29 Arnold R, Marcus JS, Petropoulos G, Schneider A. Is data the new oil? Diminishing returns to scale. 29th European Regional ITS Conference, Trento 2018; 184927, International Telecommunications Society (ITS).

30 Sharma M, Bilgic M. Evidence-based uncertainty sampling for active learning. Data Min Knowl Disc 2017; 31: 164–202.

31 Applied Logistic Regression. http://onlinelibrary.wiley.com/doi/epub/10.1002/9781118548387 (accessed September 14, 2022).

